# Pruritus and its Association with Cancer and Mortality in Dermatomyositis and Polymyositis: A Nationwide Cohort Study in Taiwan from 2005 to 2022

**DOI:** 10.1101/2024.09.26.24314441

**Authors:** Der-Jr Huang, Yu-Hsuan Joni Shao, Yi-Hsien Shih, Woan-Ruoh Lee, Ling-Ya Huang, Yu-Min Kuo, Quoc Thao Trang Pham, Hao-Jui Weng

## Abstract

**Background:** Pruritus is the most common initial symptom reported by patients with dermatomyositis and polymyositis. However, there is limited data regarding the impact of pruritus on cancer and mortality in patients with dermatomyositis and polymyositis.

**Objective:** To investigate the associations of pruritus to cancer and mortality in patients with dermatomyositis and polymyositis.

**Methods:** This nationwide, population-based retrospective cohort study included adult dermatomyositis and polymyositis patients from Taiwan’s National Health Insurance Research Database between 2005 and 2022. Sex- and age-matched pruritic patients, identified by over six weeks of antipruritic medication use, and nonpruritic patients were analyzed. The primary outcome was cancer occurrence or all-cause mortality.

**Results:** Among 919 matched pairs of pruritic and non-pruritic patients, cancer was observed in 19.96% in the long-term pruritic group, 14.63% in the short-term pruritic group, and 10.34% in the nonpruritic group (p<0.0001). All-cause mortality was documented at 30.37% in the long-term pruritic group, 29.69% in the short-term pruritic group, and 37.76% in the nonpruritic group (p<0.0001). Pruritus was associated with an increased risk of cancer (hazard ratio 1.492, 95% confidence interval 1.093-2.036), and a lower risk of all-cause mortality (hazard ratio 0.489, 95% confidence interval 0.419-0.571).

**Conclusion:** This population-based study revealed pruritus appeared to be associated with increased cancer risk and decreased all-cause mortality. Thus, pruritus may serve as a pragmatic factor for risk stratification and tailored treatment strategies in dermatomyositis and polymyositis. Comprehensive cancer screening is recommended for patients with dermatomyositis or polymyositis, particularly those presenting with pruritus, whereas patients without pruritus may require vigilant management for potentially life-threatening complications and comorbidities.

**Key points:**

1. Previous single-institutional studies and studies with small cohorts reported conflicting data regarding the impact of pruritus on cancer and mortality in patients with dermatomyositis and polymyositis
2. Pruritus in dermatomyositis and polymyositis appeared to be associated with increased cancer risk and decreased all-cause mortality.
3. Our findings suggest that pruritus may serve as a pragmatic factor for risk stratification and tailored treatment strategies in dermatomyositis and polymyositis.

## 1 Introduction

Dermatomyositis (DM) and polymyositis (PM) are rare but debilitating autoimmune diseases. In the United States, the annual incidence of DM is 9.63 cases per million people.[1] In Taiwan, DM and PM together occur at a frequency of 11.5 cases per million people.[2] Both conditions are characterized by progressive symmetrical proximal muscle weakness. DM also presents distinct skin manifestations, such as pruritus, heliotrope rash, and Gottron’s papules. Additionally, patients with DM and PM have increased risks of cancer, interstitial lung disease, stroke,[3–6] and ischemic heart disease, [5–7] and possess one of the poorest prognoses among connective tissue diseases.

Pruritus is the most common initial symptom reported by patients with DM, and associated with increased cutaneous severity. Up to 63% to 90% of patients with DM experience pruritus during the disease course.[1,8,9] Furthermore, pruritus in DM is associated with poor mental health and has a more significant impact on quality of life than other pruritic dermatosis, such as psoriasis and atopic dermatitis. [9–12]

Pruritus has been identified as a notable prognostic factor in several diseases. In conditions such as Hodgkin’s lymphoma[13,14] and hemodialysis,[15] pruritus is associated with poorer survival. Conversely, in polycythemia vera, pruritus is associated with a lower risk of arterial thrombosis and improved survival.[16] Furthermore, pruritus might serve as a marker of occult cancer, regardless of the type of underlying disease.[17] Regarding DM and PM, previous studies have shown that pruritus was correlated with an elevated risk of cancers,[18–20] while one study presented contradictory evidence.[21] This discrepancy highlights the need for large-scale studies to investigate the association between pruritus and its complications in patients with DM and PM.

In this study, we investigated the potential association between pruritus and cancer risk in patients with DM and PM, by using Taiwan’s national database and registry. We aimed to investigate the impact of pruritus on the risk of cancer in DM or PM, and the association between pruritus and overall survival in these patients. Through our investigation, we elucidated the clinical importance of pruritus, which may help stratify these patients into subgroups and guide management strategies in DM and PM.

## 2 Methods

### 2.1 Data sources

All participant data were obtained from the Registry of Catastrophic Illness Database in Taiwan, a subset of the National Health Insurance Research Database (NHIRD). This database was established for public research purposes as part of Taiwan’s National Health Insurance Program, which was initiated in 1995 and covered more than 99% of Taiwan’s 23□million residents. The case definitions of DM and PM were strictly limited to the ones who underwent comprehensive clinical and laboratory assessments that met the diagnostic criteria by Bohan and Peter.[22,23] Only patients with a diagnosis of probable or definite DM and PM were given the catastrophic illness certificate. The accuracy and validity regarding dermatomyositis and polymyositis in the Registry of Catastrophic Illness Database have been well-established in prior studies.[2,24]

### 2.2 Study design and participants

Data on patients with DM or PM diagnosed between January 1, 2005, and December 31, 2022, were extracted from the Registry of Catastrophic Illness Database in Taiwan by using the International Classification of Diseases, Ninth Revision (*ICD-9*) and International Classification of Diseases, Tenth Revision (*ICD-10*) codes (supplementary table S1). To minimize bias, patients with DM or PM with concurrent conditions potentially associated with pruritus were excluded, such as atopic dermatitis, urticaria, psoriasis, mycosis fungoides, and Sezary syndrome (as defined using *ICD-9* and *ICD-10* codes; supplementary table S1). Additionally, patients undergoing dialysis (as identified using expenditure codes; supplementary table S1) were excluded. Given the high prevalence of atopic dermatitis and urticaria, only patients with two consecutive diagnoses of these conditions were excluded.

### 2.3 Definition of pruritus

The index date for a patient with DM or PM was defined as the date of the first medical record related to the diagnosis. The cases with chronic pruritus were defined based on the methodology previously outlined by Ting et al.[25] Patients with DM or PM who were prescribed antipruritic agents for 42 consecutive days or more after the index date were included in the pruritic group (PG) (as defined using Anatomical Therapeutic Chemical codes, including systemic or topical antihistamines; menthol lotion; and combination ointments containing chlorpheniramine maleate, lidocaine hydrochloride, hexachlorophene, methyl salicylate, menthol, and camphor; supplementary table S2), and further divided into long-term pruritic group (LPG) for those treated for 84 days or more and short-term pruritic group (SPG) for those treated for 42 to 83 days. Prescriptions separated by an interval shorter than 30 days were also considered consecutive. Patients without prescriptions for antipruritic agents, or those prescribed such agents for less than 42 consecutive days, were classified into the nonpruritic group (NPG). To exclude other indications for prescribing antihistamines, prescriptions given under the diagnosis of allergic rhinitis (defined by *ICD-9* and *ICD-10* codes; supplementary table S1) were removed from the prescription records.

### 2.4 Outcome measurement

The primary outcomes of interest were cancer and ILD development, as well as mortality. Cancer development was defined as receiving a diagnosis corresponding to the International Classification of Diseases for Oncology, Third Edition codes C00–C97 in the Taiwan Cancer Registry between 2005 and 2022. The diagnoses related to nonmelanoma skin cancer and metastasis were excluded (defined by *ICD-9* and *ICD-10* codes; supplementary table S3). Cancer cases were further categorized into two groups, based on registration date within 5 years prior to, and 10 years after the index date. The mortality and cause of death were determined from 2005 to 2022, using the National Death Registry in Taiwan.

To minimize the influence of potential confounding factors, additional factors including age at index date, sex, and comorbidities known to be associated with both outcome risk and pruritus were evaluated. Comorbidities, including diabetes mellitus, hypertension, and other medical conditions were defined by *ICD-9* and *ICD-10* codes documented at least twice in outpatient department visits or once during inpatient admissions before the index date (supplementary table S1).

### 2.5 Statistical analysis

The distributions of the demographic characteristics, and comorbidities of the patients with DM and PM, were presented as frequencies (percentages) for categorical variables, medians (with interquartile ranges) for continuous variables, and means ± standard deviations for approximately normally distributed continuous variables. Differences in distribution proportions of categorical variables were assessed using the Cochran–Mantel–Haenszel test, whereas the mean differences in continuous variables between the two groups were evaluated using the generalized estimating equation. Survival analysis was performed by the Kaplan-Meier method. Hazard ratios (HRs) and 95% confidence intervals (CIs) were estimated using multivariable Cox proportional-hazard models to investigate the association between pruritus and the risk of newly diagnosed cancer development and all-cause mortality. The individual risk of each confounding factor was assessed using simple Cox proportional-hazard models, and confounders exhibiting significant outcome risks were subsequently included in multivariable Cox proportional-hazard models for adjustment. All statistical analyses were conducted using SAS software (version 9.4, SAS Institute, Cary, NC, USA), with a two-sided p-value of <0.05 considered significant.

### 2.6 Sensitivity analyses

To assess the impact of the pruritus definition cutoff point on the outcome measurement, sensitivity analyses were conducted.

## 3 Results

### Baseline characteristics of the DM/PM cohort

From 2005 to 2022, 2,723 patients listed on the NHIRD were diagnosed with DM or PM (Figure 1). Among these patients, 122 were excluded for age younger than 18 years, and 469 were excluded for concomitant pruritus-associated diseases (212 with atopic dermatitis, 95 with urticaria, 122 with psoriasis, 4 with mycosis fungoides, and 90 undergoing dialysis). A total of 2,132 patients with DM or PM remained. They were categorized into the PG (1,180 patients) and the NPG (952 patients) based on their antipruritic medication prescriptions. After matching by date of birth (±2 years) and sex at a ratio of 1:1, 919 patients of DM/PM with pruritus (PG) and 919 without (NPG) were included in the analysis.

**Fig 1.**
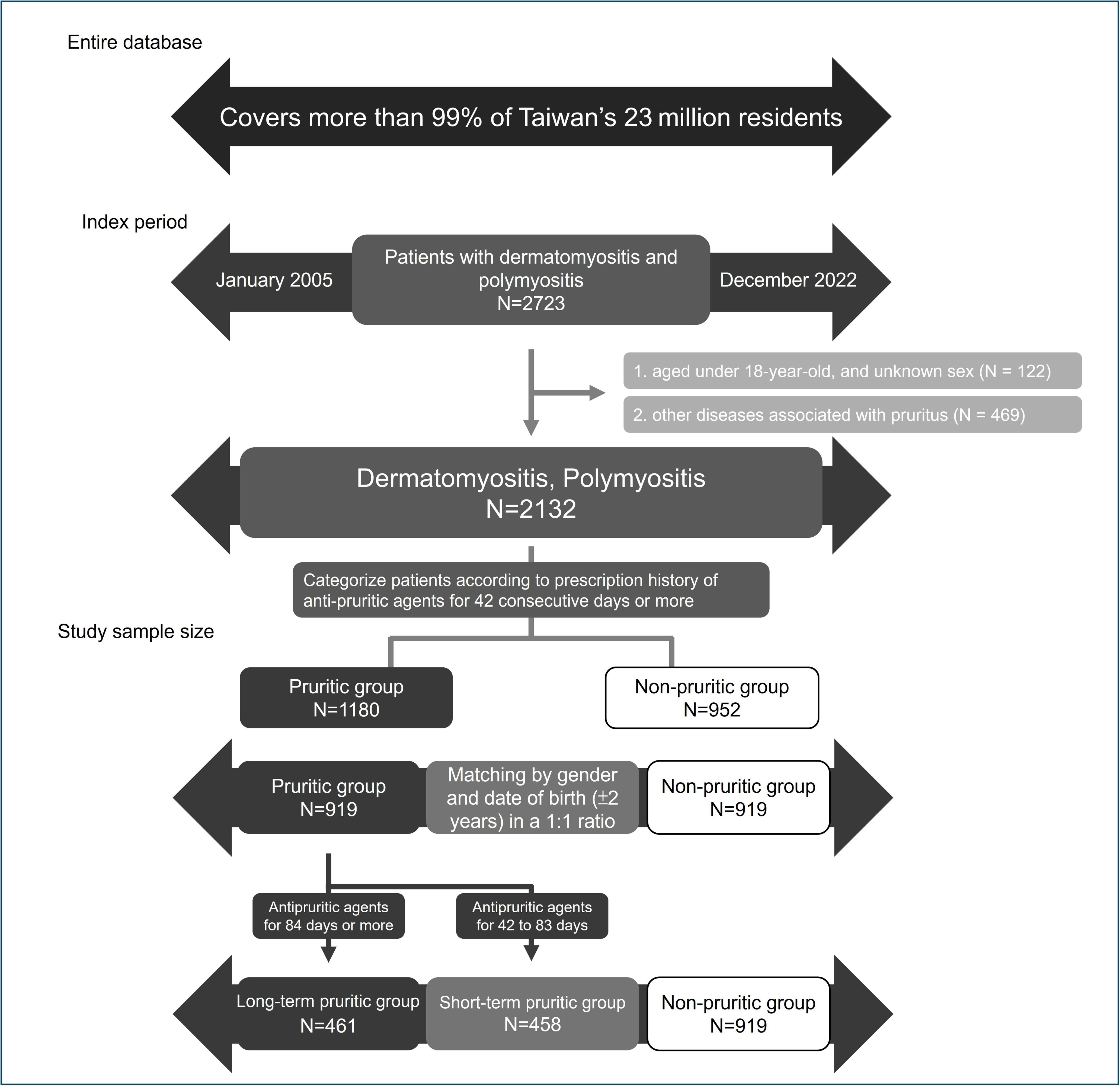
Study design. Patients with dermatomyositis and polymyositis were extracted from National Health Insurance Research Database in Taiwan from 2005 to 2022.

The demographic data of the overall cohort (patients with DM and PM) were examined. The average follow-up time was 6.7 years. Our cohort exhibited a female predominance, with a male-to-female ratio of approximately 1:2 (Table 1). The mean age at the index date was 52.0 years in the LPG, 51.6 years in the SPG, and 51.8 years in the NPG. No significant baseline difference in comorbidities was observed.

**Table 1.** Distributions of baseline characteristics in dermatomyositis and polymyositis cohort, by pruritus status, in 2005–2022.

|  | Overall cohort |  |  |  | Dermatomyositis |  |  |  | Polymyositis |  |  |  |
| --- | --- | --- | --- | --- | --- | --- | --- | --- | --- | --- | --- | --- |
|  | LPG<br>N=461 | SPG<br>N=458 | NPG<br>N=919 | p | LPG<br>N=288 | SPG<br>N=242 | NPG<br>N=427 | p | LPG<br>N=173 | SPG<br>N=216 | NPG<br>N=492 | p |
| Male, n (%) | 164<br>(35.57) | 160<br>(34.93) | 324<br>(35.26) | - | 104<br>(36.11) | 86<br>(35.54) | 147<br>(34.43) | - | 60<br>(34.68) | 74<br>(34.26) | 177<br>(35.98) | - |
| **Age at index date, year, n (%)** |  |  |  |  |  |  |  |  |  |  |  |  |
| mean (SD) | 52.0<br>(14.15) | 51.6<br>(14.91) | 51.8<br>(14.62) | 0.94 | 52.2<br>(10.01) | 50.9<br>(14.82) | 52.1<br>(14.40) | 0.32 | 51.7<br>(14.40) | 52.3<br>(15.01) | 51.5<br>(14.83) | 0.52 |
| median (IQR) | 53<br>(42-62) | 53<br>(41-62) | 52<br>(41-63) |  | 53<br>(42-62) | 51<br>(41-62) | 51<br>(42-62) |  | 53<br>(41-62) | 54<br>(43-62) | 53<br>(41-63) |  |
| min, max | 18, 88 | 18, 87 | 18, 89 |  | 18, 88 | 18, 86 | 18, 89 |  | 20, 84 | 19, 87 | 18, 87 |  |
| < 40 | 94<br>(20.39) | 94<br>(20.52) | 192<br>(20.89) | 0.76 | 57<br>(19.79) | 47<br>(19.42) | 82<br>(19.20) | 0.71 | 37<br>(21.39) | 47<br>(21.76) | 110<br>(22.36) | 0.65 |
| 40-65 | 284<br>(61.61) | 282<br>(61.57) | 557<br>(60.61) |  | 179<br>(62.15) | 155<br>(64.05) | 263<br>(61.59) |  | 105<br>(60.69) | 127<br>(58.80) | 294<br>(59.76) |  |
| > 65 | 83<br>(18.00) | 82<br>(17.90) | 170<br>(18.50) |  | 52<br>(18.06) | 40<br>(16.53) | 82<br>(19.20) |  | 31<br>(17.92) | 42<br>(19.44) | 88<br>(17.89) |  |
| **Co-morbidities, n (%)** |  |  |  |  |  |  |  |  |  |  |  |  |
| Diabetes mellitus | 62<br>(13.45) | 51<br>(11.14) | 127<br>(13.82) | 0.1 | 29<br>(13.54) | 25<br>(10.33) | 54<br>(12.65) | 0.92 | 23<br>(13.29) | 26<br>(12.04) | 73<br>(14.84) | 0.13 |
| Hypertension | 123<br>(26.68) | 119<br>(25.98) | 252<br>(27.42) | 0.54 | 74<br>(25.69) | 58<br>(23.97) | 110<br>(25.76) | 0.33 | 49<br>(28.32) | 61<br>(28.24) | 142<br>(28.86) | 0.94 |
| Vasculitis | 5<br>(1.08) | 3<br>(0.66) | 3<br>(0.33) | 0.07 | NA | NA | NA | 0.11 | NA | NA | NA | 0.32 |
| Arthritis | 22<br>(4.77) | 23<br>(5.02) | 43<br>(4.68) | 0.66 | 13<br>(4.51) | 9<br>(3.72) | 26<br>(6.09) | 0.38 | 9<br>(5.20) | 14<br>(6.48) | 17<br>(3.46) | 0.09 |

IQR, interquartile range; SD, standard deviation.
Overall cohort: patients with dermatomyositis and polymyositis.
LPG, long-term pruritic group: receiving antipruritic agents for 84 consecutive days or more.
SPG, short-term pruritic group: receiving antipruritic agents for 42 consecutive days or more, but less than 84 days.
NPG, nonpruritic group: receiving antipruritic agents for less than 42 consecutive days.

### 3.1 Association between pruritus and cancer

To investigate the association between pruritus and cancers in patients with DM or PM, we analyzed the prevalence of cancers, before and after the diagnosis of DM and PM, in the matched groups. For the overall cohort, a higher percentage of patients in the LPG and SPG developed cancer than in the NPG, over the 5 years prior to, and 10 years after the index date (Table 2; 19.96% for LPG, 14.63% for SPG, and 10.34% for NPG; p<0.0001). We next investigated the association between specific cancers and pruritus among patients with DM or PM. In our cohort, nasopharyngeal cancer, breast cancer, lung cancer, and colorectal cancer were the most prevalent cancer types, representing 20.2%, 15.4% 15.4%, and 12.5% of cancers, respectively. Increased prevalence was noted in the pruritic group for nasopharyngeal cancer (4.72% vs. 1.05%, p < 0.0001). A similar trend was observed in breast cancer (2.83% vs. 1.57%, p = 0.05).

**Table 2.** Distributions of cancer and all-cause mortality in dermatomyositis and polymyositis cohort, by pruritus status, in 2005–2022.

|  | **LPG** | **SPG** | **NPG** | **p** |
| --- | --- | --- | --- | --- |
| **Overall cohort** | **N=461** | **N=458** | **N=919** |  |
| **Cancer #** |  |  |  |  |
| Overall period | 92 (19.96) | 67 (14.63) | 95 (10.34) | 0.0002 |
| Ever registered in 5 years prior to index date | 18 (3.90) | 21 (4.59) | 32 (3.48) | 0.33 |
| Ever registered in 10 years after index date | 77 (16.70) | 50 (10.92) | 67 (7.29) | 0.0002 |
| Interstitial lung disease | 151 (32.75) | 125 (27.29) | 283 (30.79) | 0.39 |
| All-cause mortality | 140 (30.37) | 136 (29.69) | 347 (37.76) | <0.0001 |
| Cancer-associated mortality | 53 (11.50) | 31 (6.77) | 54 (5.88) | 0.04 |
| **Dermatomyositis** | **N=288** | **N=242** | **N=427** |  |
| **Cancer #** |  |  |  |  |
| Overall period | 75 (26.04) | 47 (19.42) | 57 (13.35) | 0.11 |
| Ever registered in 5 years prior to index date | 13 (4.51) | 17 (7.02) | 23 (5.39) | 0.43 |
| Ever registered in 10 years after index date | 63 (21.88) | 34 (14.05) | 38 (8.90) | 0.12 |
| Interstitial lung disease | 90 (31.25) | 75 (30.99) | 168 (39.34) | 0.39 |
| All-cause mortality | 95 (32.99) | 76 (31.40) | 199 (46.60) | 0.0007 |
| Cancer-associated mortality | 46 (15.97) | 22 (9.09) | 34 (7.96) | 0.3 |
| **Polymyositis** | **N=173** | **N=216** | **N=492** |  |
| **Cancer #** |  |  |  |  |
| Overall period | 17 (9.83) | 20 (9.26) | 38 (7.72) | 0.35 |
| Ever registered in 5 years prior to index date | 5 (2.89) | 4 (1.85) | 9 (1.83) | 0.39 |
| Ever registered in 10 years after index date | 14 (8.09) | 16 (7.41) | 29 (5.89) | 0.48 |
| Interstitial lung disease | 61 (35.26) | 50 (23.15) | 115 (23.37) | 0.24 |
| All-cause mortality | 45 (26.01) | 60 (27.78) | 148 (30.08) | 0.03 |
| Cancer-associated mortality | 7 (4.05) | 9 (4.17) | 20 (4.07) | 1 |

Data are presented as *n* (%).
Overall cohort: patients with dermatomyositis and polymyositis.
LPG, long-term pruritic group: receiving antipruritic agents for 84 consecutive days or more.
SPG, short-term pruritic group: receiving antipruritic agents for 42 consecutive days or more, but less than 84 days.
NPG, nonpruritic group: receiving antipruritic agents for less than 42 consecutive days.

### 3.2 Association between pruritus and mortality

In contrast to the association observed between cancer and pruritus, we identified a significantly lower all-cause mortality rate in the LPG and the SPG, than in the NPG (Table 2). Among the overall cohort during the 18-year follow-up period, the all-cause mortality rate was 30.37% in the LPG, 29.69% in the SPG, and 37.76% in the NPG (p<0.0001). Further analysis of the causes of death in the study population, revealed an elevated risk of cancer-related mortality in the LPG and SPG, among patients with DM and PM. For the overall cohort, our findings revealed 11.50% of patients in the LPG, and 6.77% of patients in the SPG, died from various types of cancers, whereas only 5.88% died in the NPG (p=0.04).

Kaplan-Meier survival curves for the overall cohort (Figure 2) showed a significant difference between the PG and the NPG (p<0.0001). A higher proportion of deaths in the NPG occurred within the first year after DM or PM diagnosis, compared to deaths in the PG. Among the overall cohort, the 1-year survival rate was 95% in the PG, compared to only 77% in the NPG. This disparity was particularly pronounced among patients with DM, where the 1-year survival rate was 93% in the PG, compared to 68% in the NPG.

**Fig 2.**
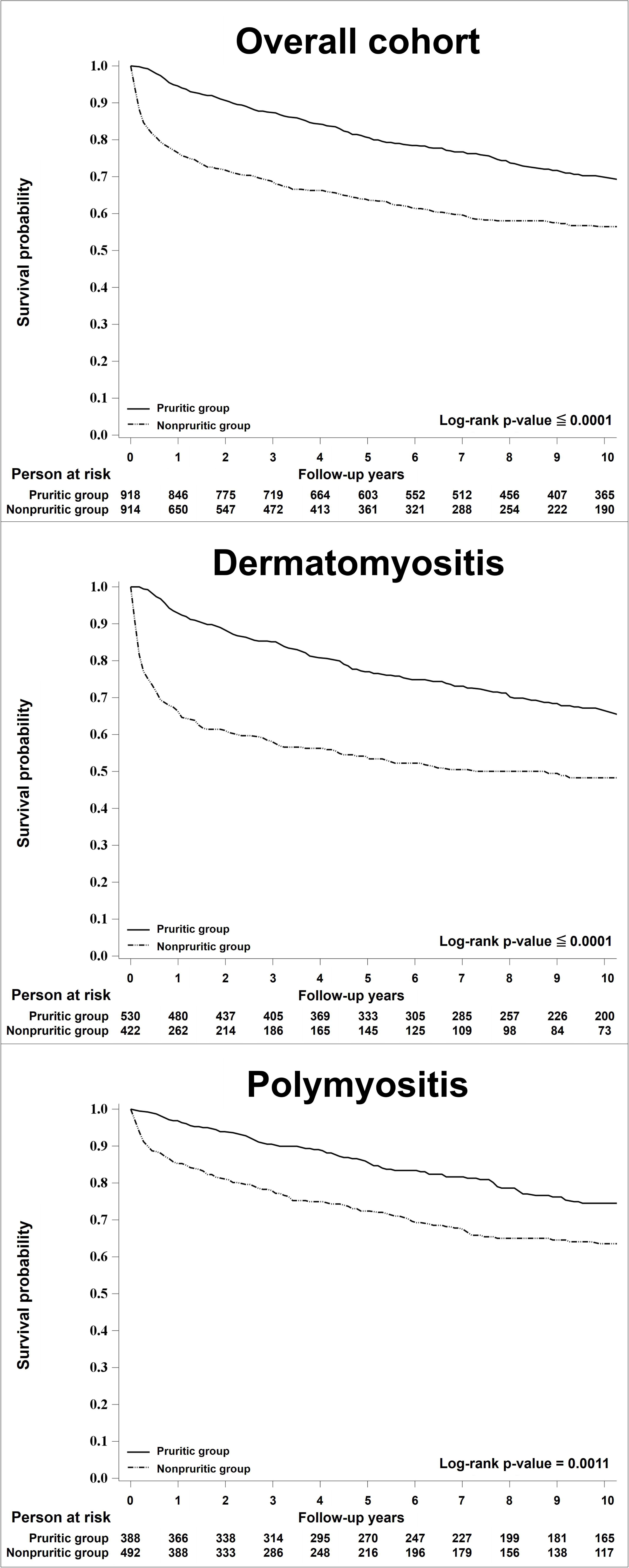
Survival curves. Upper, overall cohort (dermatomyositis and polymyositis); middle, dermatomyositis; lower, polymyositis.

### 3.3 Hazard ratio analysis of pruritus in DM and PM

To investigate the variables for patients’ survival, simple Cox proportional-hazard models were utilized. The results supported our previous data, that the presence of pruritus during the disease course, was associated with a higher risk of newly diagnosed cancers for the overall cohort (supplementary table S5; HR 1.561, 95% CI 1.154-2.111). Subgroup analysis revealed that long-term pruritus was significantly associated with cancer (HR 1.971, 95% CI 1.416-2.744), while short-term pruritus showed no significant association with cancer. Furthermore, older age of onset, including ages between 40 and 65 years, >65 years, male sex, diabetes mellitus, hypertension, and vasculitis, were significantly associated with newly diagnosed cancers.

The presence of pruritus, in contrast, was negatively associated with the mortality for the overall cohort (supplementary table S6; HR 0.564, 95% CI 0.490-0.650). Subgroup analysis identified both long-term and short-term pruritus were significantly associated with mortality (HR 0.591, 95% CI 0.494-0.707; HR 0.539, 95% CI 0.450-0.646, respectively). However, older age of onset, including an age between 40 and 65 years, >65 years, male sex, diabetes mellitus, hypertension, ILD, and cancer, were positively associated with mortality.

In the multivariable analysis for newly diagnosed cancer, adjustment was performed for significant confounding factors, including sex, age, diabetes, hypertension, vasculitis, and arthritis. Additionally, ILD and cancer were considered in the analysis of mortality risk. The multivariate analysis (Table 3) indicated that pruritus was associated with an increased risk of developing cancer (HR 1.492, 95% CI 1.093-2.036), with a more pronounced effect for long-term pruritus (HR 1.849, 95% CI 1.314-2.602). Conversely, pruritus was associated with a lower risk of mortality (HR 0.489, 95% CI 0.419-0.571).

**Table 3.** Event risk in dermatomyositis cohort, 2005-2022.

|  | Adjusted hazard ratio (95% confidence interval) |  |
| --- | --- | --- |
|  | Newly diagnosed cancer | All-cause mortality |
| **Overall** |  |  |
| Pruritus (Yes vs. No) | **1.492 (1.093-2.036)** | **0.489 (0.419-0.571)** |
| Pruritus (vs. No) |  |  |
| SPG | 1.139 (0.775-1.674) | **0.468 (0.385-0.568)** |
| LPG | **1.849 (1.314-2.602)** | **0.511 (0.424-0.617)** |
| **Dermatomyositis** |  |  |
| Pruritus (Yes vs. No) | **1.707 (1.126-2.587)** | **0.413 (0.335-0.509)** |
| Pruritus (vs. No) |  |  |
| SPG | 1.220 (0.733-2.029) | **0.379 (0.291-0.495)** |
| LPG | **2.107 (1.360-3.266)** | **0.444 (0.349-0.564)** |
| **Polymyositis** |  |  |
| Pruritus (Yes vs. No) | 0.874 (0.523-1.461) | **0.515 (0.402-0.660)** |
| Pruritus (vs. No) |  |  |
| SPG | 0.889 (0.482-1.639) | **0.536 (0.396-0.726)** |
| LPG | 0.855 (0.443-1.652) | **0.490 (0.355-0.675)** |

Data are presented as hazard ratio (95% confidence interval).
Overall cohort: patients with dermatomyositis and polymyositis.
LPG, long-term pruritic group: receiving antipruritic agents for 84 consecutive days or more.
SPG, short-term pruritic group: receiving antipruritic agents for 42 consecutive days or more, but less than 84 days.
NPG, nonpruritic group: receiving antipruritic agents for less than 42 consecutive days.
Post cancer model of patients without baseline cancer adjusted for sex, age, diabetes, hypertension, vasculitis(overall); sex, age, diabetes, hypertension (dermatomyositis); age, hypertension (polymyositis).
Post Interstitial lung disease model of patients without baseline cancer adjusted for sex, age, arthritis (overall, dermatomyositis); age (polymyositis).
All-cause death model adjusted for sex, age, diabetes, hypertension, arthritis, cancer, interstitial lung disease (overall); sex, age, diabetes, hypertension, cancer, interstitial lung disease (dermatomyositis, polymyositis).

The sensitivity analyses confirmed the robustness of the results (supplementary table S7). We consistently observed associations between pruritus and cancer, as well as pruritus and mortality, across various pruritus duration thresholds.

## 4 Discussion

This study demonstrated the association between pruritus in patients with DM and PM, and their comorbidities and prognostic outcomes at a national level. Patients with DM and PM experiencing pruritus were found to have a higher risk of developing cancers, but a lower risk of all-cause mortality, compared to nonpruritic patients. This finding was complemented by similar results in the sensitivity analyses. More than half of our cohort experienced chronic pruritus, highlighting the need for ongoing and comprehensive screening for cancers in this high-risk patient population.

Pruritus has long been documented in autoimmune connective tissue diseases.[26] Approximately 57% of patients with autoimmune connective tissue diseases, irrespective of their specific diagnoses, report pruritus upon their initial presentation.[27] The prevalence of pruritus varies across different diseases, ranging from 42% to 62% in systemic sclerosis,[27–30] 38% to 53% in Sjögren syndrome,[31,32] and 63% to 90% in DM.[1,8,9] Pruritus in lupus erythematosus (LE) is generally considered less severe than that in DM.[12,27] Studies have indicated that the median pruritus visual analogue scale score in LE is significantly lower than that observed in DM (3.8 vs. 2.0).[12] Moreover, the majority of patients with LE experience only mild (45% to 62%) or moderate (54%) pruritus,[27,33] which contrasts with reports from patients with DM. For the underlying cause, small fiber neuropathies, paraneoplastic symptoms, and involvement of IL-31 are believed to contribute to DM-associated itch. Kim et al. revealed upregulation of IL-31 and its receptor alpha gene in the lesional skin of patients with DM,[9] with IL-31 exhibiting a significant positive correlation with the severity of itch.

Assessing antihistamine usage may serve as a surrogate indicator for a patient’s pruritus in our study as previously reported.[25] In clinical practice, sedating antihistamines are commonly prescribed as first-line treatment for pruritus.[34] They alleviate pruritus by blocking H1 receptors on C-afferent fiber terminals,[35] preventing mast cell degranulation,[36,37] and providing a soporific effect. While not all pruritic diseases could be effectively controlled by antihistamines, most patients with chronic pruritus, regardless of its cause, will receive H1 antihistamines at some point to alleviate the itch.[38,39] Antihistamines are favored for their proven efficacy in other pruritic dermatosis, such as chronic idiopathic urticaria,[40–43] atopic dermatitis, and psoriasis,[44] as well as their affordability, accessibility, and minimal side effects. The cut-off point for antihistamine usage was set at 6 weeks in our study, as chronic pruritus is defined by the International Forum for the Study of Itch as itching that persists for over 6 weeks.[45] Short-term antihistamine uses for other conditions such as allergic reactions were excluded.

Using multivariable Cox proportional-hazard models, we found that DM and PM patients experiencing pruritus exhibited a significantly higher risk of newly diagnosed cancer (Table 3; adjusted HR 1.492, 95% CI 1.093-2.036). Previous studies show that DM patients with pruritus may be predisposed to internal malignancies,[18–20] with one contradictory study of 63 patients in Germany.[21] However, due to limited sample sizes in earlier studies, their findings regarding the association between pruritus and cancer, appeared inconclusive or underpowered. Our findings, in contrast, provided robust evidence supporting the notion that DM and PM patients with pruritus exhibit an elevated risk of cancer.

Many autoimmune diseases have been associated with a higher risk of developing cancers. For example, Sjögren’s syndrome is associated with an increased risk of non-Hodgkin lymphoma,[46,47] and systemic lupus erythematosus is associated with non-Hodgkin lymphoma and lung cancer.[48–50] Additionally, systemic sclerosis is linked to an elevated risk of lung cancer.[51–53] DM is associated with a higher cancer risk compared with other autoimmune diseases. Approximately 2.2% to 5% of patients with Sjögren’s syndrome develop cancers,[47,54,55] compared to a higher prevalence of 9.4% to 32.3% among patients with DM.[2,56–60] Some scholars have postulated that in DM, cancer itself triggered autoimmunity rather than as the result of disordered immune function or immunosuppressive drugs, which have been implicated in the pathogenesis of cancer in other autoimmune diseases.[61–64] In our analysis, the majority of the patients with DM or PM developed cancers within a year before or after the diagnosis of DM or PM. This is consistent with the findings of prior research. Furthermore, studies have indicated that the increased risk of cancers may persist beyond the initial 5-year period. Thus, sustained and comprehensive cancer screening is necessary for patients with DM or PM, particularly those deemed to be at high risk following their diagnosis.

The multivariable Cox proportional-hazard models also identified pruritus as a protective factor against all-cause mortality in patients with DM and PM (Table 3; adjusted HR 0.489, 95% CI 0.419-0.571). Prior studies showed cancer, ILD, infection, and cardiac involvement as predominant causes of death in DM and PM patients.[65–73] The most common cancer associated with DM in East Asia is nasopharyngeal carcinoma, however, it is less associated with immediate patient mortality. In contrast, ILD, particularly rapidly progressive interstitial lung disease, is associated with high mortality rates within the first year of diagnosis[74,75], thus, frequently necessitating aggressive immunosuppressive therapy in ILD patients.[76–79] Further analysis of death causes, revealed a higher incidence of deaths from infection in the non-pruritic group (NPG), within the first year of DM or PM diagnosis. This supports the concept that non-pruritic patients may require higher doses of steroids and immunosuppressants, to manage their disease.

The survival curve indicated a marked decline in survival during the first year in the NPG, a pattern not observed in the pruritic group (PG). Previous studies illustrated that lung complications are the major cause of death within the first 12 months, following DM and PM diagnosis.[80,81] This disparity in survival between the PG and NPG was even more pronounced among DM patients. It is well-documented that DM is associated with a poorer prognosis compared to PM, particularly concerning ILD[82] and cancer-related mortality.[83,84] However, although no significant difference in ILD prevalence was observed between the PG and NPG, the clinical course of ILD may vary considerably. Rapidly progressive ILD is associated with significantly poorer prognosis, compared to chronic ILD. This form of ILD is more prevalent in DM than in other connective tissue diseases,[85] and has been extensively reported in Asia, including Japan, Hong Kong, and Taiwan.[86–89]

Several hypotheses may explain the paradoxical finding of higher cancer risks but lower all-cause mortality rates in our pruritic DM and PM patients. First, disease severity in the nonpruritic group might be higher than in the pruritic group and required a higher dosage of immunosuppressant. We observed that more patients in the nonpruritic group died from infection, which was consistent with our hypothesis that nonpruritic patients require higher dosages of immunosuppressants to control their disease. Secondly, the nonpruritic group might consist of a higher prevalence of overlap syndrome. Researchers have categorized DM into two groups: pure DM and overlap myositis with dermatomyositis features (OMDM). [90] Pure DM is associated with higher cancer risks but an excellent 15-year survival rate of 92%. By contrast, patients with OMDM often develop myositis before cutaneous symptoms, and possess a decreased 15-year survival rate of 65%. Thirdly, distinct myositis-specific and myositis-associated autoantibodies might be involved in the pathogenesis of pruritic and nonpruritic patients. Several autoantibodies may be candidates for pruritic and nonpruritic conditions. Anti-transcriptional intermediary factor 1 γ (anti-TIF1γ) antibodies and anti-small ubiquitin-like modifier-1 activating enzyme (anti-SAE) antibodies are often associated with extensive skin involvement and a higher risk of cancer.[91–95] Conversely, patients with anti-melanoma differentiation-associated protein 5 (anti-MDA5) antibodies and anti-signal recognition particle (anti-SRP) antibodies typically present with less pruritic skin lesions, and a poor prognosis, despite having a lower risk of cancer.[91,96,97]Further investigation is warranted to prove our hypotheses.

This study has several limitations. First, because our analysis relied on data from an administrative database, some potential ascertainment problems were unavoidable, including coding errors and mortality misclassifications. However, differences in coding error between the pruritic and non-pruritic groups are unlikely. Additionally, our study benefited from the stringent regulation of Taiwan’s Registry of Catastrophic Illness Database, which requires DM or PM cases to meet the criteria set by Bohan and Peter.[22,23] Secondly, although assessing pruritus through prescription medication is an objective and quantitative approach, it inevitably underestimates patients with mild pruritus who did not require a prescription, those who discontinued antipruritic medication due to non-responsiveness, and those who used over-the-counter medications or traditional Chinese herbal treatments for pruritus. Thirdly, we were unable to analyze clinical parameters such as cutaneous manifestations other than pruritus, laboratory parameters, and myositis-associated or specific antibodies because of the inherent limitations of the NHIRD. Finally, given that the population data were derived from a single country (Taiwan), particularly the significant association of dermatomyositis to nasopharyngeal cancer, further research is required to assess the generalizability and reproducibility of these findings in other populations.

## 5 Conclusions

This study employed nationwide insurance administration data, and revealed a significant association between pruritus and cancer, as well as cancer-related mortality, in patients with DM and PM. Thus, comprehensive cancer screening is recommended for patients with DM or PM, particularly those presenting with pruritus. Notably, nonpruritic patients were associated with a higher risk of all-cause mortality, especially during the first year after diagnosis. Therefore, patients without pruritus may require vigilant management for potentially life-threatening complications and comorbidities.

## Declarations

### Consent to participate (including appropriate statements)

Not applicable

### Ethical approval

The study was approved by the Joint Institutional Review Board of Taipei Medical University (N202403054). The requirement for informed consent was waived by the Institutional Review Board because the dataset was deidentified.

### Prior presentation

The abstract of this study was presented at EADV Congress 2024 at Amsterdam on 26 September 2024.

### Reprint requests

None

### Data Availability Statement

Data are available from the National Health Insurance Research Database (NHIRD) published by the Taiwan National Health Insurance (NHI) Bureau. Due to legal restrictions imposed by the government of Taiwan in relation to the Personal Information Protection Act, data cannot be made publicly available. Requests for data can be sent as a formal proposal to the NHIRD (https://nhird.nhri.org.tw).

## Acknowledgments

We acknowledge the statistical support of the Health Data Analytics and Statistics Center, Office of Data Science, Taipei Medical University, Taiwan. This work was supported by Taipei Medical University-Shuang Ho Hospital, Ministry of Health and Welfare (112YSR-02), and National Science and Technology Council (110-2314-B-038-028-MY3).

## Author contributions

DJH conceptualised the study, contributed to the study design and interpretation of the results, and drafted the manuscript. YHJS contributed to the study design, data extraction, and analysis, and reviewed the manuscript. WRL and YHS contributed to the study design and reviewed the manuscript. LYH contributed to the study design, data analysis, and interpretation of the results, and reviewed the manuscript. YMK and QTTP contributed to the interpretation of the results and reviewed the manuscript. HJW conceptualized the study, contributed to the study design and interpretation of the results, and reviewed the manuscript. All authors had access to the final study results and accepted responsibility for submitting them for publication. The corresponding author attests that all listed authors meet authorship criteria and that no others meeting the criteria have been omitted. HJW is the guarantor.

## Abbreviations

DM: dermatomyositis
PM: polymyositis
ILD: interstitial lung disease
NHIRD: National Health Insurance Research Database
International Classification of Diseases, Ninth Revision: *ICD-9*
International Classification of Diseases, Tenth Revision: *ICD-10*
HR: hazard ratio
CI: confidence interval
PG: pruritic group
LPG: long-term pruritic group
SPG: short-term pruritic group
NPG: nonpruritic group

