## supplementary tables for "Pruritus and its Association with Cancer and Mortality in Dermatomyositis and Polymyositis: A Nationwide Cohort Study in Taiwan from 2005 to 2022"

**Supplementary Table S1. Study sample**

| Drug/disease | ATC/ICD/expenditure codes | Definition |
| --- | --- | --- |
| Inclusion |  |  |
| Other dermatomyositis | *ICD-9*: 710.3  *ICD-10*: M33.1–M33.19 | Catastrophic Illness Database |
| Polymyositis | *ICD-9*: 710.4  *ICD-10*: M33.2–M33.29 | Catastrophic Illness Database |
| Dermatopolymyositis | *ICD-9*: 710.3  *ICD-10*: M33.9–M33.99 | Catastrophic Illness Database |
| Exclusion |  |  |
| Atopic dermatitis | *ICD-9*: 691.8  *ICD-10*: L20–L30 | Two consecutive diagnostic codes |
| Urticaria | *ICD-9*: 708  *ICD-10*: L50.9 | Two consecutive diagnostic codes |
| Psoriasis | *ICD-9*: 696.1  *ICD-10*: L40–L40.9 | Diagnostic code assigned at outpatient, inpatient, and emergency departments |
| Mycosis fungoides | *ICD-9*: 202.1  *ICD-10*: C84.0 |  |
| Sezary disease | *ICD-9*: 202.2  *ICD-10*: C84.10 |  |
| Dialysis | 58001C, 58002C, 58019C, 58020C, 58021C, 58022C, 58023C, 58024C, 58025C, 58027C, 58029C |  |
| Allergic rhinitis | *ICD-9*: 477.0, 477.1, 477.8, 477.9, 472.0, 472.1, 472.2  *ICD-10*: J30, J31 |  |
| Comorbidities | | |
| Diabetes mellitus | *ICD-9*: 250  *ICD-10*: E08–E13 | Diagnostic code assigned at outpatient, inpatient, and emergency departments |
| Hypertension | *ICD-9*: 401–405  *ICD-10*: I10–I15 |  |
| Vasculitis | *ICD-9*: 709.1, 695.89  *ICD-10*: L95 |  |
| Arthritis | *ICD-9*: 716.20–716.68  *ICD-10*: M13 |  |

**Supplementary Table S2. Antipruritic agents**

| Drug/disease | ATC/*ICD* code |
| --- | --- |
| Antihistamines for systemic use | R06 |
| Antihistamines for topical use | D04AA |
| Menthol lotion | D04AX |
| CB strong  (combination ointments containing chlorpheniramine maleate, lidocaine hydrochloride, hexachlorophene, methyl salicylate, menthol, and camphor) | D04AB01 |

**Supplementary Table S3. Outcome measurements**

| Endpoints | *ICD-9* and *ICD-10* codes and *ICD-O-3* | *definition* |
| --- | --- | --- |
| All types of cancer | All types of cancer: *ICD-O-3* (C00–C97)  Excluded  Nonmelanoma skin cancer (*ICD-9*: 173–173.9; *ICD-O*: C44–C44.99),  Secondary cancer (*ICD-9*: 196–199; *ICD-O*: C7B.00–C7B.8, C77.0–C77.9, C78.0–C78.89, C79.00–C79.9, R18.0, J91.0, C80.0),  Lymph node metastasis (*ICD-9*: 195–195.9; *ICD-O*: C76–C76.8) | Taiwan Cancer Registry |
| Specific cancer | The lips, oral cavity, or pharynx (*ICD-9*: 141, 145, 146, 148; *ICD-O*: C02, C06, C09, C10, C13)  Nasopharynx (*ICD-9*: 147; *ICD-O*: C11)  Esophagus (*ICD-9*: 150; *ICD-O*: C15)  Stomach (*ICD-9*: 151; *ICD-O*: C16)  Colon (*ICD-9*: 153; *ICD-O*: C18)  Rectum (*ICD-9*: 154; *ICD-O*: C20)  The liver and intrahepatic bile ducts (*ICD-9*: 155; *ICD-O*: C22)  Other or unspecified parts of the biliary tract (*ICD-9*: 156; *ICD-O*: C24)  The larynx (*ICD-9*: 161; *ICD-O*: C32)  The bronchus and lung (*ICD-9*: 162; *ICD-O*: C34)  Secondary and unspecified malignant neoplasm of lymph nodes or leukemia (*ICD-9*: 169, 196; *ICD-O*: C42, C77)  Breast (*ICD-9*: 174; *ICD-O*: C50)  Corpus uteri (*ICD-9*: 182; *ICD-O*: C54)  Ovary (*ICD-9*: 183; *ICD-O*: C56)  Prostate (*ICD-9*: 185; *ICD-O*: C61)  Bladder (*ICD-9*: 188; *ICD-O*: C67) | Taiwan Cancer Registry |
| Interstitial lung disease | *ICD-9*: 515, 516.3, 516.8, 516.9  *ICD-10*: J84 | Diagnostic code assigned at outpatient twice, or inpatient once |

**Supplementary Table S4. Duration of antipruritic medication use.**

|  | **PG** | **NPG** |
| --- | --- | --- |
|  | N=919 | N=919 |
| mean (std) | 558.1 (759.72)^*^ | 7.3 (7.45)^#^ |
| median (IQR) | 266 (104-660)^*^ | 6.0 (0.0-11.3)^#^ |

IQR, interquartile range; SD, standard deviation.

PG, pruritic group: receiving antipruritic agents for 42 consecutive days or more .

NPG, nonpruritic group: receiving antipruritic agents for less than 42 consecutive days.

^*^ Total cumulative days of continuous use of antipruritic medication meeting the definition of pruritus.

^#^ Continuous days of antipruritic medication use not meeting the definition of pruritus.

**Supplementary Table S5. Risk of newly diagnosed cancer in dermatomyositis and polymyositis cohort without baseline cancer in 2005–2022.**

|  | Crude hazard ratio (95% confidence interval) | | |
| --- | --- | --- | --- |
|  | Overall cohort | Dermatomyositis | Polymyositis |
| Pruritus (Yes vs. No) | **1.561 (1.154-2.111)** | **1.744 (1.161-2.620)** | 0.939 (0.561-1.572) |
| Pruritus (vs. No) |  |  |  |
| < 84 | 1.167 (0.798-1.706) | 1.215 (0.734-2.009) | 0.955 (0.517-1.765) |
| 84+ | **1.971 (1.416-2.744)** | **2.205 (1.434-3.389)** | 0.919 (0.477-1.774) |
| Male vs. female | **1.666 (1.242-2.236)** | **1.434 (1.021-2.014)** | **2.298 (1.372-3.850)** |
| Age at index date, year |  |  |  |
| < 40 | 1.0 | 1.0 | 1.0 |
| 40-65 | **2.677 (1.634-4.384)** | **2.342 (1.305-4.204)** | **3.368 (1.320-8.596)** |
| > 65 | **4.123 (2.377-7.150)** | **3.769 (1.977-7.187)** | **5.033 (1.768-14.330)** |
| Co-morbidities (Yes vs. No) |  |  |  |
| Diabetes mellitus | **1.589 (1.086-2.326)** | **1.852 (1.202-2.855)** | 1.179 (0.543-2.560) |
| Hypertension | **2.044 (1.517-2.755)** | **2.019 (1.395-2.923)** | **2.418 (1.415-4.133)** |
| Vasculitis | **3.582 (1.057-12.138)** | 1.939 (0.470-7.994) | 10.487 (0.741-148.416) |
| Arthritis | 0.549 (0.227-1.327) | 0.404 (0.129-1.266) | 0.951 (0.234-3.867) |

Data are presented as hazard ratio (95% confidence interval).

**Supplementary Table S6. All-cause mortality risk in dermatomyositis and polymyositis cohort in 2005–2022.**

|  | Crude hazard ratio (95% confidence interval) | | |
| --- | --- | --- | --- |
|  | Overall cohort | Dermatomyositis | Polymyositis |
| Pruritus (Yes vs. No) | **0.564 (0.490-0.650)** | **0.455 (0.375-0.552)** | **0.659 (0.524-0.829)** |
| Pruritus (vs. No) |  |  |  |
| < 84 | **0.539 (0.450-0.646)** | **0.425 (0.330-0.549)** | **0.663 (0.500-0.879)** |
| 84+ | **0.591 (0.494-0.707)** | **0.481 (0.383-0.603)** | **0.653 (0.482-0.885)** |
| Male vs. female | **1.512 (1.280-1.786)** | **1.381 (1.121-1.703)** | **1.695 (1.309-2.195)** |
| Age at index date, year |  |  |  |
| < 40 | 1.0 | 1.0 | 1.0 |
| 40-65 | **2.867 (2.090-3.933)** | **2.449 (1.675-3.580)** | **3.568 (2.043-6.230)** |
| > 65 | **9.010 (6.521-12.449)** | **6.523 (4.395-9.680)** | **14.342 (8.211-25.050)** |
| Co-morbidities (Yes vs. No) |  |  |  |
| Diabetes mellitus | **1.918 (1.574-2.337)** | **1.626 (1.239-2.133)** | **2.489 (1.867-3.318)** |
| Hypertension | **2.389 (2.022-2.823)** | **2.065 (1.653-2.580)** | **3.050 (2.382-3.905)** |
| Vasculitis | 0.644 (0.166-2.501) | 0.353 (0.056-2.240) | 1.512 (0.174-13.111) |
| Arthritis | **0.588 (0.354-0.977)** | 0.530 (0.278-1.013) | 0.659 (0.290-1.495) |
| Cancer | **2.750 (2.097-3.608)** | **2.185 (1.613-2.960)** | **4.247 (2.370-7.612)** |
| Interstitial lung disease | **1.897 (1.548-2.325)** | **1.597 (1.200-2.126)** | **2.430 (1.835-3.216)** |

Data are presented as hazard ratio (95% confidence interval).

**Supplementary Table S7 Sensitivity analysis, Event risk in dermatomyositis cohort, 2005-2022.**

|  | Adjusted hazard ratio (95% confidence interval) | |
| --- | --- | --- |
| Pruritus (Yes vs. No) | **Newly diagnosed cancer** | **All-cause mortality** |
| Overall |  |  |
| 28-days | **1.446 (1.026-2.040)** | **0.400 (0.335-0.478)** |
| 56-days | **1.849 (1.384-2.470)** | **0.574 (0.491-0.672)** |
| 84-days | **1.441 (1.077-1.919)** | **0.541 (0.455-0.643)** |
| Dermatopolymyositis and other dermatomyositis |  |  |
| 28-days | **1.662 (1.066-2.590)** | **0.351 (0.280-0.441)** |
| 56-days | **2.293 (1.556-3.380)** | **0.480 (0.392-0.587)** |
| 84-days | **1.486 (1.030-2.143)** | **0.463 (0.371-0.578)** |
| Polymyositis |  |  |
| 28-days | 0.807 (0.450-1.448) | **0.381 (0.285-0.509)** |
| 56-days | 0.910 (0.558-1.482) | **0.623 (0.482-0.804)** |
| 84-days | 0.903 (0.537-1.521) | **0.539 (0.405-0.717)** |

Data are presented as n (%).

**28-days**

Post cancer model of patients without baseline cancer adjusted for sex, age, diabetes, hypertension, vasculitis (overall); age, diabetes, hypertension (dermatopolymyositis and other dermatomyositis); sex, age, hypertension, vasculitis (polymyositis).

Post Interstitial lung disease model of patients without baseline cancer adjusted for vasculitis (overall).

All-cause death model adjusted for sex, age, diabetes, hypertension, cancer (overall, dermatopolymyositis and other dermatomyositis, polymyositis).

**56-days**

Post cancer model of patients without baseline cancer adjusted for sex, age, diabetes, hypertension (overall); age, diabetes, hypertension (dermatopolymyositis and other dermatomyositis); sex, age, hypertension, vasculitis (polymyositis).

Post Interstitial lung disease model of patients without baseline cancer adjusted for age (dermatopolymyositis and other dermatomyositis).

All-cause death model adjusted for sex, age, diabetes, hypertension, arthritis, cancer (overall, dermatopolymyositis and other dermatomyositis, polymyositis).

**84-days**

Post cancer model of patients without baseline cancer adjusted for sex, age, diabetes, hypertension, vasculitis (overall); age, diabetes, hypertension (dermatopolymyositis and other dermatomyositis); sex, age, hypertension, vasculitis (polymyositis).

Post Interstitial lung disease model of patients without baseline cancer adjusted for vasculitis (overall); age, hypertension (dermatopolymyositis and other dermatomyositis).

All-cause death model adjusted for sex, age, diabetes, hypertension, arthritis, cancer (overall, dermatopolymyositis and other dermatomyositis); sex, age, diabetes, hypertension, cancer (polymyositis).
